# Influenza hospitalization burden by subtype, age, comorbidity and vaccination status: 2012/13 to 2018/19 seasons, Quebec, Canada

**DOI:** 10.1101/2023.08.04.23293392

**Authors:** Sara Carazo, Charles-Antoine Guay, Danuta M Skowronski, Rachid Amini, Hugues Charest, Gaston De Serres, Rodica Gilca

## Abstract

**Background:** The primary objective of influenza immunization programs is to reduce the risk and burden of severe outcomes. To inform optimal program strategies, we monitored influenza hospitalizations over several seasons of varying subtype predominance, stratified by age, comorbidity and vaccination status.

**Methods:** We assembled data from an active hospital-based surveillance network involving systematic swabbing and PCR-confirmation of influenza virus infection by type/subtype during peak-weeks of seven influenza seasons (2012/13 to 2018/19) in Quebec, Canada. We estimated seasonal, population-based incidence of influenza-associated hospitalizations (interpreted as risk) by subtype, age, comorbidity and vaccine status, and derived the number-needed-to-vaccinate to prevent one hospitalization per stratum.

**Results:** The average seasonal incidence of influenza-associated hospitalization was 89/100,000 (95%CI: 86, 93), lower during A(H1N1) (49-82/100,000) than A(H3N2) seasons (73-143/100,000). Overall risk followed a J-shaped age pattern, highest among infants 0-5 months and adults ≥75 years. Hospitalization risks were highest for children <5 years during A(H1N1) but for adults ≥75 years during A(H3N2) subtype- predominant seasons. Age-adjusted hospitalization risks were 7-fold higher among individuals with versus without comorbidities (214 versus 30/100,000). The number-needed-to-vaccinate to prevent hospitalization was 82-fold lower for ≥75-years-olds with comorbidity (n=1,995), who comprised 39% of all hospitalizations, than for healthy 18-64-year-olds (n=163,488), who comprised just 6% of all hospitalizations.

**Conclusions:** In the context of broad-based influenza immunization programs (targeted or universal), severe outcome risks should be simultaneously examined by subtype, age, comorbidity, and vaccine status. Policymakers require such detail to prioritize further promotional efforts and expenditures toward the greatest and most efficient program impact.

**40-word summary:** This hospital-based study involving systematic PCR testing over seven seasons revealed important differences in influenza hospitalization risk by subtype, age, comorbidity, and vaccination status. The findings highlight the need for data-driven decision-making to optimize vaccination strategies and minimize healthcare burden.

## INTRODUCTION

Seasonal influenza remains one of the leading causes of morbidity and mortality globally, despite broad-based vaccination efforts. In the United States (US) alone, seasonal influenza accounts for >$3 billion in direct annual health care costs, most of which is attributable to the nearly 400,000 hospitalizations that accrue on average annually(1). The primary goal of most influenza vaccination programs is to reduce the risk and burden due to severe outcomes (hospitalizations and deaths). Even in the context of high vaccine coverage among target groups and universal influenza immunization programs throughout much of North America (including the US and most provinces of Canada), estimates of the residual hospitalization burden due to influenza are seldom examined by vaccination status(2–4).

Like most countries of Europe, the influenza immunization program in the province of Quebec, Canada, is targeted toward those at highest risk of severe outcome, notably elderly adults and those ≥ 6 months of agewith underlying comorbidity(5,6). All jurisdictions, however, struggle to understand which strategies may have the greatest and most efficient impact in further reducing the residual hospitalization burden (e.g. existing targeted with enhanced promotion, expanded targeted and/or universal). To explore this question, we assembled data from an active hospital–based respiratory virus surveillance network and examined influenza-associated hospitalisation risks stratified simultaneously by age, comorbidity, and vaccination status during seven influenza seasons (2012-13 to 2018-19) of varying type/subtype predominance.

## METHODS

### 1. Hospital-based surveillance

The hospital–based surveillance network in the province of Quebec included two community and two academic/tertiary acute-care regional hospitals, serving nearly 10% of the overall Quebec population (8,3M in 2018)(7). Patients were recruited during influenza epidemic peak-weeks. The latter were defined by percent positivity reported by a sentinel laboratory surveillance network in Quebec comprised of >40 hospital laboratories that test >100,000 respiratory specimens for influenza virus, on average per year. The peak-week period began when influenza positivity was ≥15% for two consecutive weeks and ended when positivity first fell <15% or when the budgeted sample size was exhausted (∼800- 1000 specimens per season) (**Supplementary Figure 1**).

During these seasonal peak-week periods, all patients presenting to participating hospital emergency departments with an acute respiratory infection (ARI: fever/feverishness in absence of an identifiable non-respiratory cause, or cough, or sore throat) were systematically swabbed. Patients admitted for ≥24 hours were invited to participate in the study. Demographic and clinical details were collected on consenting patients by research nurses through patient (or legal representative) interview and chart review. Vaccination status was based on self-reported receipt of seasonal influenza vaccine at least 14 days before symptom onset. Patients missed by the nurse because of early discharge or who refused or were unable to participate still contributed to surveillance with their laboratory results aggregated by age category.

Nasal specimens collected on flocked swabs were tested by the multiplex Luminex® RVP FAST version 1 or the NxTAG RPP assays which detects influenza A (seasonal H3N2 and H1N1 subtypes), influenza B, and 15 other respiratory viruses(8). The two assays are approved for diagnosis by Health Canada authorities.

### 2. Estimated hospitalizations

Influenza-associated hospitalizations were estimated (stratified by age group, comorbidity and/or season) with adjustment for varying peak-week contribution per season, test sensitivity and percentage participation (**Supplementary Figure 1, Supplementary Table 1**). To account for varying peak-week contribution to the overall seasonal span, we derived the peak-week influenza detections as a percentage of all influenza detections across the season by the sentinel laboratory surveillance network (53% to 73%). We multiplied the inverse of that seasonal peak-week percent contribution by the peak-week tally of hospitalizations in order to estimate the number of hospitalizations for each full season.

Hospitalization may be delayed in relation to symptom onset and test sensitivity is reduced among patients consulting >8 days post onset(9). We therefore applied the influenza positivity of patients consulting ≤8 days post-onset to the tally of patients consulting >8 days post-onset (or with unknown onset date) whose multiplex results for respiratory viruses were otherwise fully negative. Finally, percentage participation (i.e. included divided by eligible individuals) ranged 59-87% by both age group and season and estimated hospitalization incidences were also adjusted for that variation.

### 3. Population denominators

Age-specific denominators for the underlying catchment area of participating hospitals were obtained from the *Institut de la statistique du Québec*(10). Age-specific denominators further stratified for comorbidity were estimated based upon respondents to a population-based vaccination survey conducted in Quebec in 2018, using the percentage that reported select comorbid conditions known to increase the risk of influenza complications, including 34%, 45% and 59% of adults 18-64, 65-74 and ≥75- years old, respectively(11). Since the survey data aggregated all adults ≥75 years, the proportion with comorbidity among age groups 75-84 and ≥85-year-olds was estimated by applying the coefficient of increase in the prevalence of multimorbidity by age reported in Quebec in 2016-17(12). For children, the proportion with comorbidities was based on that reported by controls randomly sampled from the general Quebec population as part of previous case-control studies, including 3%, 7% and 6% for 6-23 month-, 2-4-year- and 5-17-year-olds, respectively(13,14). Because comorbidity estimates were not available for 0-5-month-olds, this group was not further stratified on that basis. Finally, age- (18-64, 65- 74 and ≥75 years) and comorbidity-specific denominators were further stratified on vaccination status using average coverages extracted from the 2016 and 2018 influenza vaccine coverage surveys for the province of Quebec(11,15).

### 4. Estimated hospitalization incidence/risk and number-needed-to-vaccinate (NNV)

Influenza hospitalization incidences (interpreted as risks) per 100,000 population were estimated as the adjusted number of hospitalizations divided by corresponding denominators. Estimates were stratified by season (2012-13 to 2018-19), age group (0-5 and 6-23 months, 2-4, 5-17, 18-64, 65-74, 75-84 and ≥85 years), comorbidity status (≥1 or no comorbid condition) and vaccination status. Mean incidences were calculated using a 1000 bootstrap re-sampling with replacement method. The 95% confidence intervals (CI) were defined as the 2.5^th^ and 97.5^th^ percentiles of the bootstrap distribution.

The NNV to prevent one hospitalization was estimated as per below, for vaccine effectiveness (VE) of 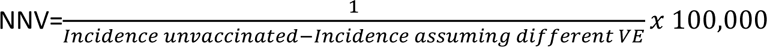

### 5. Ethics

For the first three years, participating hospitals each obtained Institutional Review Board approval from Comité d’éthique de la recherche du Centre hospitalier de l’Université Laval, from Comité d’éthique de la recherche du Centre de santé et des services sociaux de Chicoutimi, from Comité scientifique et d’éthique de la recherche du Centre de santé et de services sociaux de Laval, from Comité central d’éthique de la recherche du ministre de la Santé et des Services sociaux du Centre de santé et de services sociaux du Haut-Richelieu et Centre de santé et de services sociaux de Rimouski-Neigette.

Thereafter, activities were incorporated into routine provincial surveillance, mandated by the Ministry of Health, and a waiver was obtained from the Research Ethics Board of the Centre hospitalier universitaire de Québec-Université Laval.

## RESULTS

The seven influenza seasons of the study included four influenza A(H3N2) predominant seasons (2012- 13, 2014-15, 2016-17 and 2017-18) and three A(H1N1) predominant seasons (2013-14, 2015-16 and 2018-19) (**Supplementary Figure 1**). Influenza B often circulated after seasonal peak weeks and was therefore less reliably captured by the hospital surveillance network.

### Population

Among the 6,545 eligible patients admitted with ARI, 4,821 (74%) consented to participate. The main reasons for non-inclusion were: missed by the study nurse (44%), unable to consent (32%), sampling or laboratory problem (8%) and refusal (6%) (**Supplementary Table 2**). Non-participants were older, more often infected with A(H3N2), and previously recruited in earlier seasons (not shown). Among the 1,171 and 3,650 hospitalized children and adults who agreed to participate, 272 (23%) and 1,682 (46%), respectively, tested influenza-positive with characteristics shown in **Table 1**. Age distribution varied substantially by influenza A subtype. Children and adults <60 years were more affected during A(H1N1)- predominant seasons, comprising 56% of hospitalizations compared to only 27% during A(H3N2) seasons. In contrast, adults ≥75 years old were disproportionately affected during A(H3N2)-predominant seasons, comprising 56% of hospitalizations compared to 26% during A(H1N1)-predominant seasons.

**Table 1.** Characteristics of patients hospitalized with influenza by season of influenza A(H3N2) or A(H1N1) predominance.

|  | A(H3N2) seasons<sup>a</sup> | A(H1N1) seasons<sup>b</sup> | Total |
| --- | --- | --- | --- |
|  | N (%) | N (%) | N (%) |
| Total | 1136 | 540 | 1954 |
| Age |  |  |  |
| 0-5m | 34 (2.7) | 16 (2.3) | 50 (2.6) |
| 6-23m | 37 (2.9) | 60 (8.7) | 97 (5.0) |
| 2-4y | 32 (2.5) | 37 (5.4) | 69 (3.5) |
| 5-17y | 32 (2.5) | 24 (3.5) | 56 (2.9) |
| 18-64y | 207 (16.4) | 249 (36.1) | 456 (23.3) |
| 65-74y | 218 (17.2) | 124 (18.0) | 342 (17.5) |
| 75-84y | 369 (29.2) | 114 (16.55) | 483 (24.7) |
| 85+y | 336 (26.6) | 65 (9.43) | 401 (20.5) |
| Sex |  |  |  |
| Male | 568 (44.9) | 312 (45.3) | 880 (45.1) |
| Female | 697 (55.1) | 377 (54.7) | 1074 (55.0) |
| Comorbidity status |  |  |  |
| Healthy | 266 (21.0) | 235 (34.1) | 501 (25.6) |
| Cardiac comorbidity only | 311 (24.6) | 102 (14.8) | 413 (21.1) |
| Respiratory comorbidity only | 244 (19.3) | 165 (24.0) | 409 (20.9) |
| Cardiac and respiratory | 161 (12.7) | 105 (15.2) | 266 (13.6) |
| Other comorbidities<sup>c</sup> | 283 (22.4) | 82 (11.9) | 365 (18.7) |
| Vaccination status |  |  |  |
| Yes | 478 (37.8) | 196 (28.5) | 674 (34.5) |
| No | 609 (48.1) | 474 (68.8) | 1083 (55.4) |
| Unknown | 178 (14.1) | 19 (2.8) | 197 (10.1) |
| Origin of patients<sup>d</sup> |  |  |  |
| Community | 943 (74.6) | 583 (84.7) | 1526 (78.1) |
| Long term care facility | 157 (12.4) | 26 (3.8) | 183 (9.4) |
| Other facilities / institutions<sup>e</sup> | 165 (13.0) | 77 (11.2) | 242 (12.4) |
| Type of infection |  |  |  |
| Influenza only | 1099 (86.9) | 601 (87.2) | 1700 (87.0) |
| Influenza+RSV | 34 (2.7) | 14 (2.0) | 48 (2.5) |
| Influenza+ORV<sup>f</sup> | 132 (10.4) | 74 (10.7) | 206 (10.5) |
| Interval (in days) between symptom onset and sampling, mean ± standard deviation | 4.6 ± 5.6 | 3.9 ± 4.6 | 4.3 ± 5.3 |

<sup>a</sup>2012/13, 2014/15, 2016/17 and 2017/18 seasons
<sup>b</sup>2013/14, 2015/16 and 2018/19 seasons
<sup>c</sup>Obesity, diabetes, immunopathy, nephropathy, neuropathy
<sup>d</sup>Two missing values for origin of admission in H1N1 seasons
<sup>e</sup>Including intermediate care and family-type facilities, etc.
<sup>f</sup>Including human metapneumovirus, human parainfluenza viruses, adenovirus, entero/rhinovirus (not differentiated) and human bocavirus
Abbreviations: ORV, other respiratory viruses; RSV, respiratory syncytial virus;

Adults 65-74 years represented 17-18% of all hospitalizations regardless of subtype. Comorbidity was reported overall by 74% of hospitalized cases: 55% had at least one chronic cardiac and/or respiratory condition and 19% reported another chronic condition (**Table 1**). Overall, 12% of hospitalized children >6 months old and 47% of hospitalized adults had received influenza vaccine, higher among children and adults with (23% and 51%, respectively) than without (9% and 27%, respectively) comorbidities (**Figure 1**). The distribution of specific comorbidities by age group and the percent vaccinated by each is presented in **Supplementary Table 3**.

**Figure 1.**
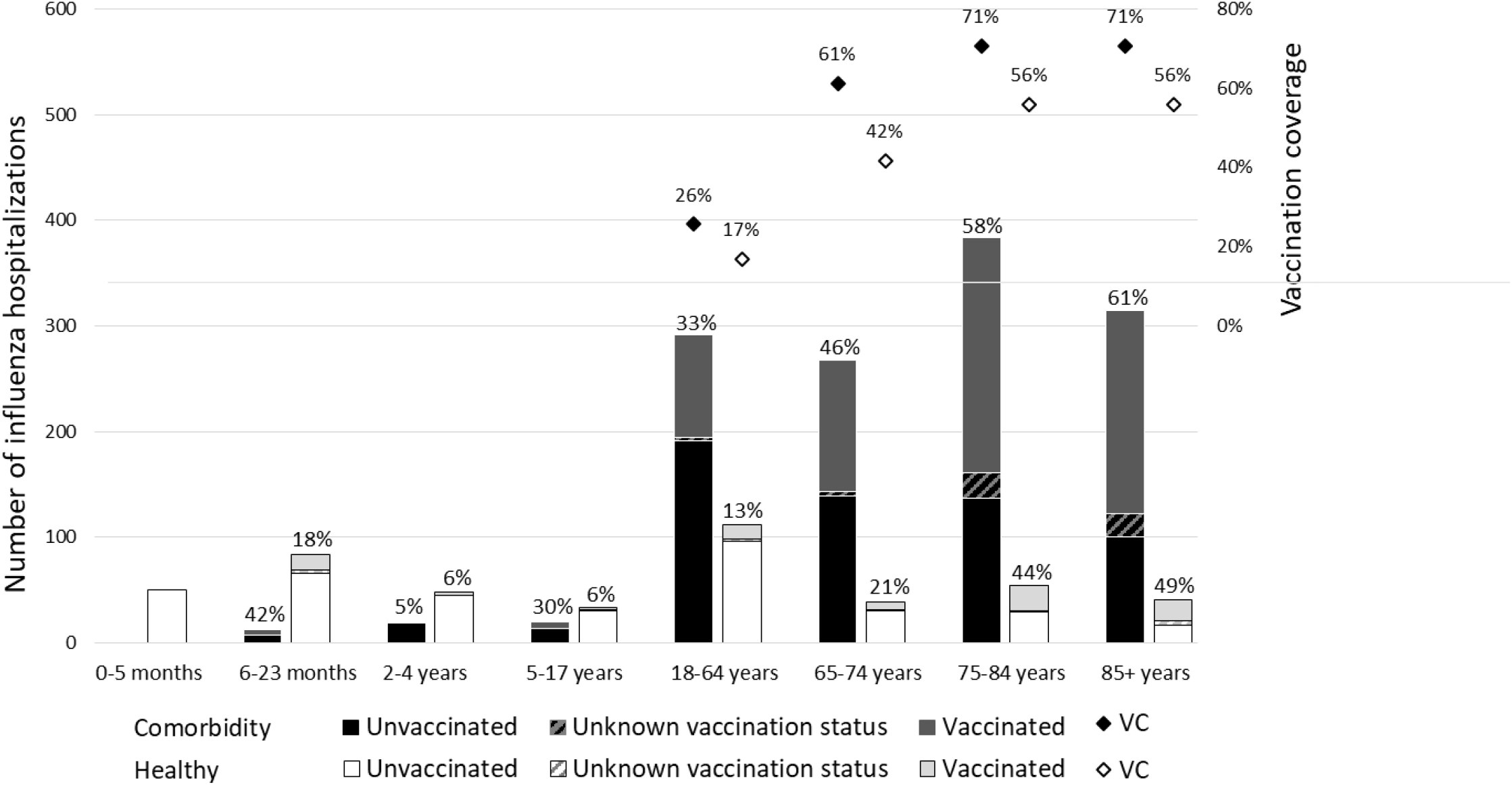
Number of influenza hospitalizations in the Quebec hospital surveillance (2012-13 to 2018-19) by age group, comorbidity status and vaccination status and vaccination coverage among adults by age group and comorbidity status Notes: Numbers over the columns are percentage of vaccinated among each age and risk subgroup (also represented by the upper grey area of each column) Vaccination coverages (VC) are the average of 2016 and 2018 Quebec’s vaccination surveys. Estimations are representative for population with and without comorbidities for 3 age groups (18-64 years, 65-74 years and ≥75 years)

### Influenza-associated hospitalization incidence risk

In the source population of about 135,000 children and 560,000 adults, the average seasonal incidence of influenza-associated hospitalizations was 89/100,000 (95%CI: 86, 93), lower during A(H1N1) predominance (lowest in 2013-14, 49/100,000; 95%CI: 42, 56) and higher during A(H3N2) predominance (highest in 2014-15, 143/100,000; 95%CI: 130, 158) (**Table 2**, **Figure 2**). The average annual hospitalization risk overall was 7-fold higher among patients with (214/100,000; 95%CI: 204, 223) versus without comorbidities (30/100,000; 95%CI: 27, 32), with similar trend in all age groups and for both influenza A subtypes.

**Figure 2.**
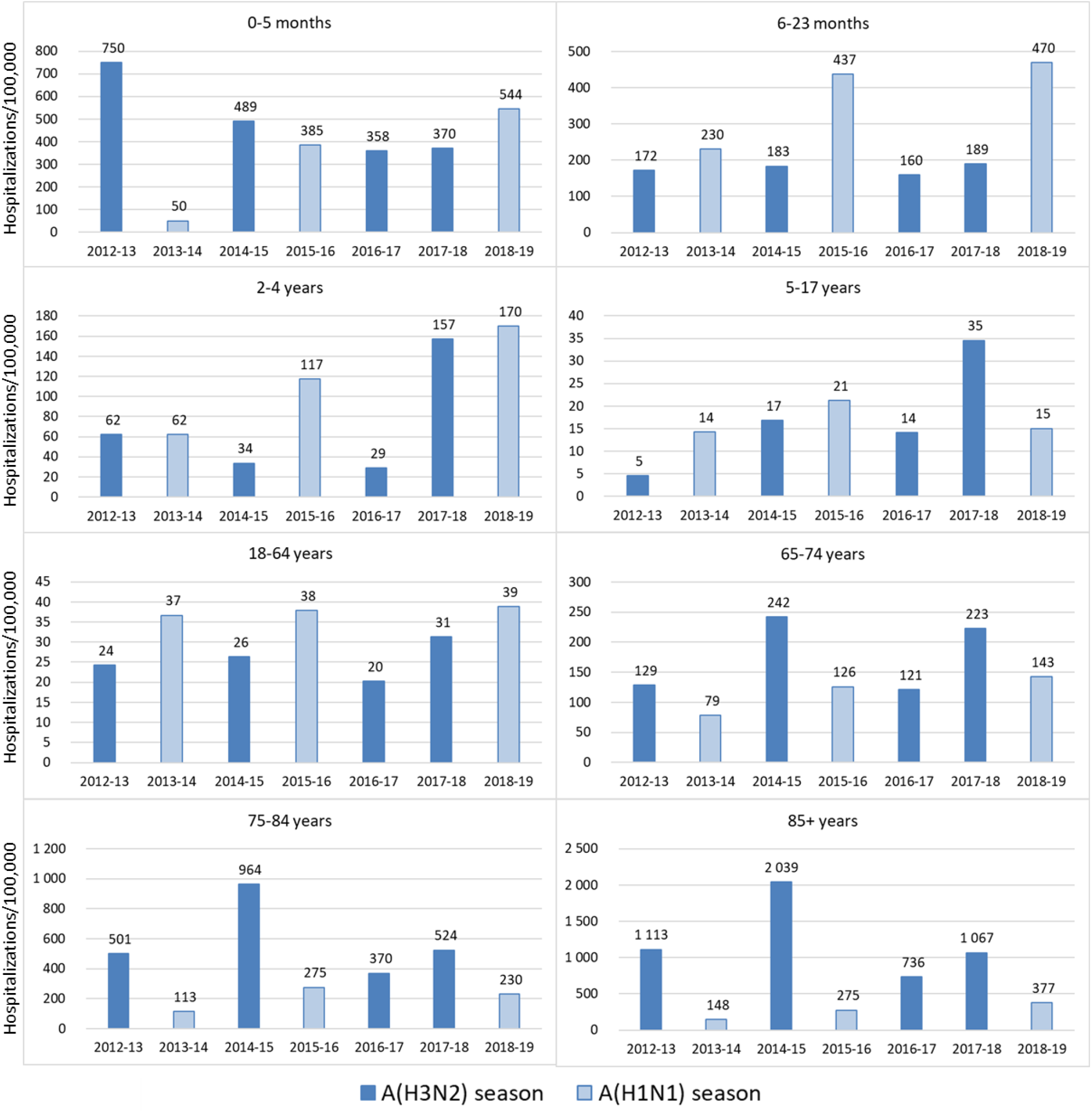
Estimated influenza-associated hospitalization rates (per 100,000) by age group and by influenza sub-type dominance season

**Table 2.**
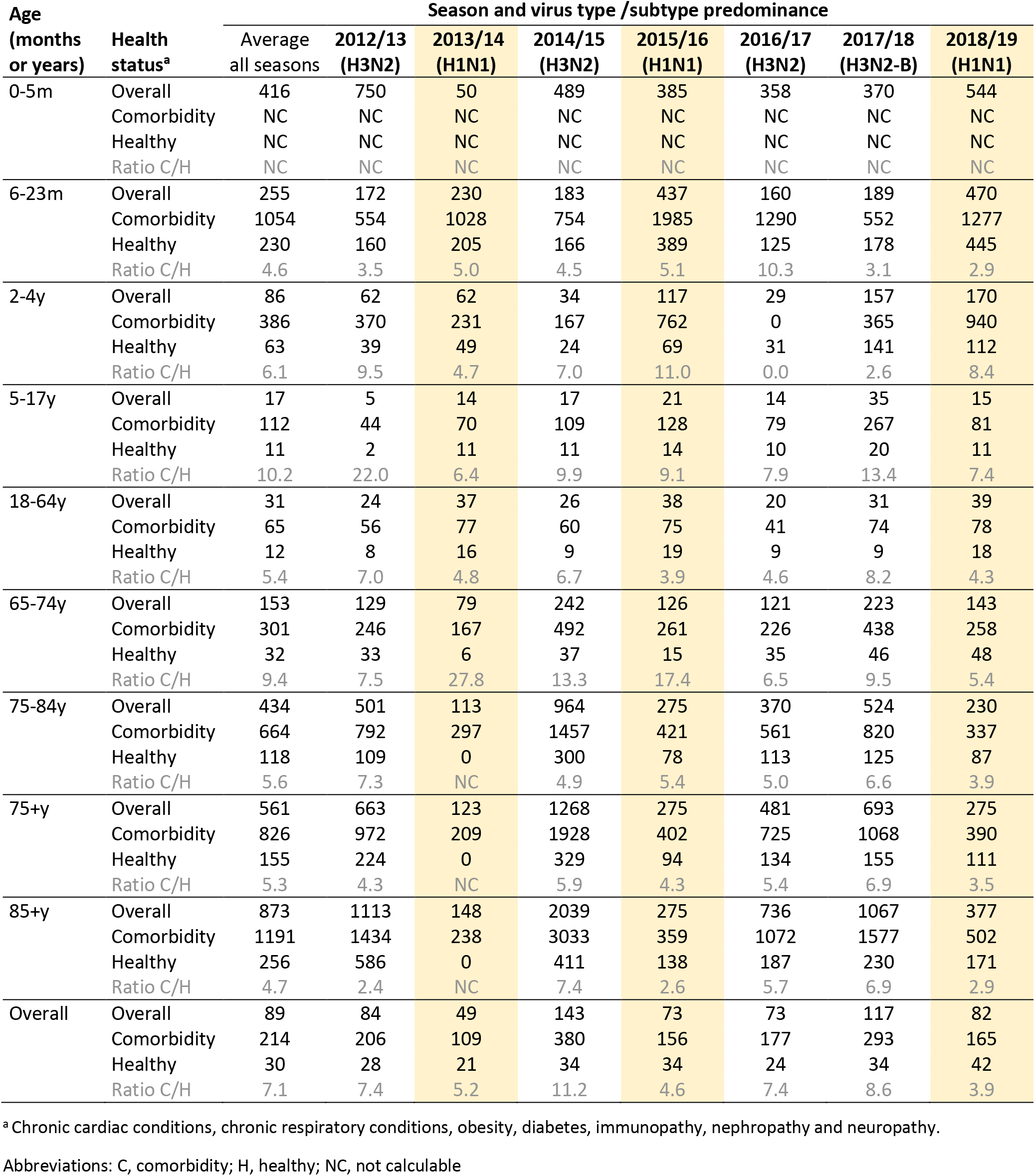
Estimated influenza-associated hospitalization incidence (per 100,000), by age group and comorbidity status, Quebec hospital surveillance from 2012-13 to 2018-19.

| Age<br>(months<br>or years) | Health<br>status<sup>a</sup> | Season and virus type /subtype predominance |  |  |  |  |  |  |  |
| --- | --- | --- | --- | --- | --- | --- | --- | --- | --- |
|  |  | Average<br>all seasons | 2012/13<br>(H3N2) | 2013/14<br>(H1N1) | 2014/15<br>(H3N2) | 2015/16<br>(H1N1) | 2016/17<br>(H3N2) | 2017/18<br>(H3N2-B) | 2018/19<br>(H1N1) |
| 0-5m | Overall | 416 | 750 | 50 | 489 | 385 | 358 | 370 | 544 |
|  | Comorbidity | NC | NC | NC | NC | NC | NC | NC | NC |
|  | Healthy | NC | NC | NC | NC | NC | NC | NC | NC |
|  | Ratio C/H | NC | NC | NC | NC | NC | NC | NC | NC |
| 6-23m | Overall | 255 | 172 | 230 | 183 | 437 | 160 | 189 | 470 |
|  | Comorbidity | 1054 | 554 | 1028 | 754 | 1985 | 1290 | 552 | 1277 |
|  | Healthy | 230 | 160 | 205 | 166 | 389 | 125 | 178 | 445 |
|  | Ratio C/H | 4.6 | 3.5 | 5.0 | 4.5 | 5.1 | 10.3 | 3.1 | 2.9 |
| 2-4y | Overall | 86 | 62 | 62 | 34 | 117 | 29 | 157 | 170 |
|  | Comorbidity | 386 | 370 | 231 | 167 | 762 | 0 | 365 | 940 |
|  | Healthy | 63 | 39 | 49 | 24 | 69 | 31 | 141 | 112 |
|  | Ratio C/H | 6.1 | 9.5 | 4.7 | 7.0 | 11.0 | 0.0 | 2.6 | 8.4 |
| 5-17y | Overall | 17 | 5 | 14 | 17 | 21 | 14 | 35 | 15 |
|  | Comorbidity | 112 | 44 | 70 | 109 | 128 | 79 | 267 | 81 |
|  | Healthy | 11 | 2 | 11 | 11 | 14 | 10 | 20 | 11 |
|  | Ratio C/H | 10.2 | 22.0 | 6.4 | 9.9 | 9.1 | 7.9 | 13.4 | 7.4 |
| 18-64y | Overall | 31 | 24 | 37 | 26 | 38 | 20 | 31 | 39 |
|  | Comorbidity | 65 | 56 | 77 | 60 | 75 | 41 | 74 | 78 |
|  | Healthy | 12 | 8 | 16 | 9 | 19 | 9 | 9 | 18 |
|  | Ratio C/H | 5.4 | 7.0 | 4.8 | 6.7 | 3.9 | 4.6 | 8.2 | 4.3 |
| 65-74y | Overall | 153 | 129 | 79 | 242 | 126 | 121 | 223 | 143 |
|  | Comorbidity | 301 | 246 | 167 | 492 | 261 | 226 | 438 | 258 |
|  | Healthy | 32 | 33 | 6 | 37 | 15 | 35 | 46 | 48 |
|  | Ratio C/H | 9.4 | 7.5 | 27.8 | 13.3 | 17.4 | 6.5 | 9.5 | 5.4 |
| 75-84y | Overall | 434 | 501 | 113 | 964 | 275 | 370 | 524 | 230 |
|  | Comorbidity | 664 | 792 | 297 | 1457 | 421 | 561 | 820 | 337 |
|  | Healthy | 118 | 109 | 0 | 300 | 78 | 113 | 125 | 87 |
|  | Ratio C/H | 5.6 | 7.3 | NC | 4.9 | 5.4 | 5.0 | 6.6 | 3.9 |
| 75+y | Overall | 561 | 663 | 123 | 1268 | 275 | 481 | 693 | 275 |
|  | Comorbidity | 826 | 972 | 209 | 1928 | 402 | 725 | 1068 | 390 |
|  | Healthy | 155 | 224 | 0 | 329 | 94 | 134 | 155 | 111 |
|  | Ratio C/H | 5.3 | 4.3 | NC | 5.9 | 4.3 | 5.4 | 6.9 | 3.5 |
| 85+y | Overall | 873 | 1113 | 148 | 2039 | 275 | 736 | 1067 | 377 |
|  | Comorbidity | 1191 | 1434 | 238 | 3033 | 359 | 1072 | 1577 | 502 |
|  | Healthy | 256 | 586 | 0 | 411 | 138 | 187 | 230 | 171 |
|  | Ratio C/H | 4.7 | 2.4 | NC | 7.4 | 2.6 | 5.7 | 6.9 | 2.9 |
| Overall | Overall | 89 | 84 | 49 | 143 | 73 | 73 | 117 | 82 |
|  | Comorbidity | 214 | 206 | 109 | 380 | 156 | 177 | 293 | 165 |
|  | Healthy | 30 | 28 | 21 | 34 | 34 | 24 | 34 | 42 |
|  | Ratio C/H | 7.1 | 7.4 | 5.2 | 11.2 | 4.6 | 7.4 | 8.6 | 3.9 |

<sup>a</sup> Chronic cardiac conditions, chronic respiratory conditions, obesity, diabetes, immunopathy, nephropathy and neuropathy.
Abbreviations: C, comorbidity; H, healthy; NC, not calculable

Influenza hospitalization risk followed a J-shaped pattern by age: 416/100,000 (95%CI: 308, 531) among 0-5-month-olds and 255/100,000 (95%CI: 204, 306) among 6-23-month-olds, lower in other pediatric age groups and increasing through adulthood, highest at 434/100,000 (95%CI: 392, 485) among 75-84-year-olds and 873/100,000 (95%CI: 751, 1019) among ≥85-year-olds. Patients 5-64 years had the lowest incidence of hospitalization (17 to 31/100,000). Compared with children 2–4 years of age, the risk of hospitalization was 5-fold higher for infants 0-5 months and 3-fold higher for children 6-23 months (**Table 2**, **Figure 3**). Compared with 65-74-year-olds, the risk of hospitalization was 3-fold higher for 75- 84-year-olds and 6-fold higher for ≥85-year-olds. The risk of hospitalization was higher among 18-64- year-olds with comorbidity than among healthy older adults 65-74 years old.

**Figure 3.**
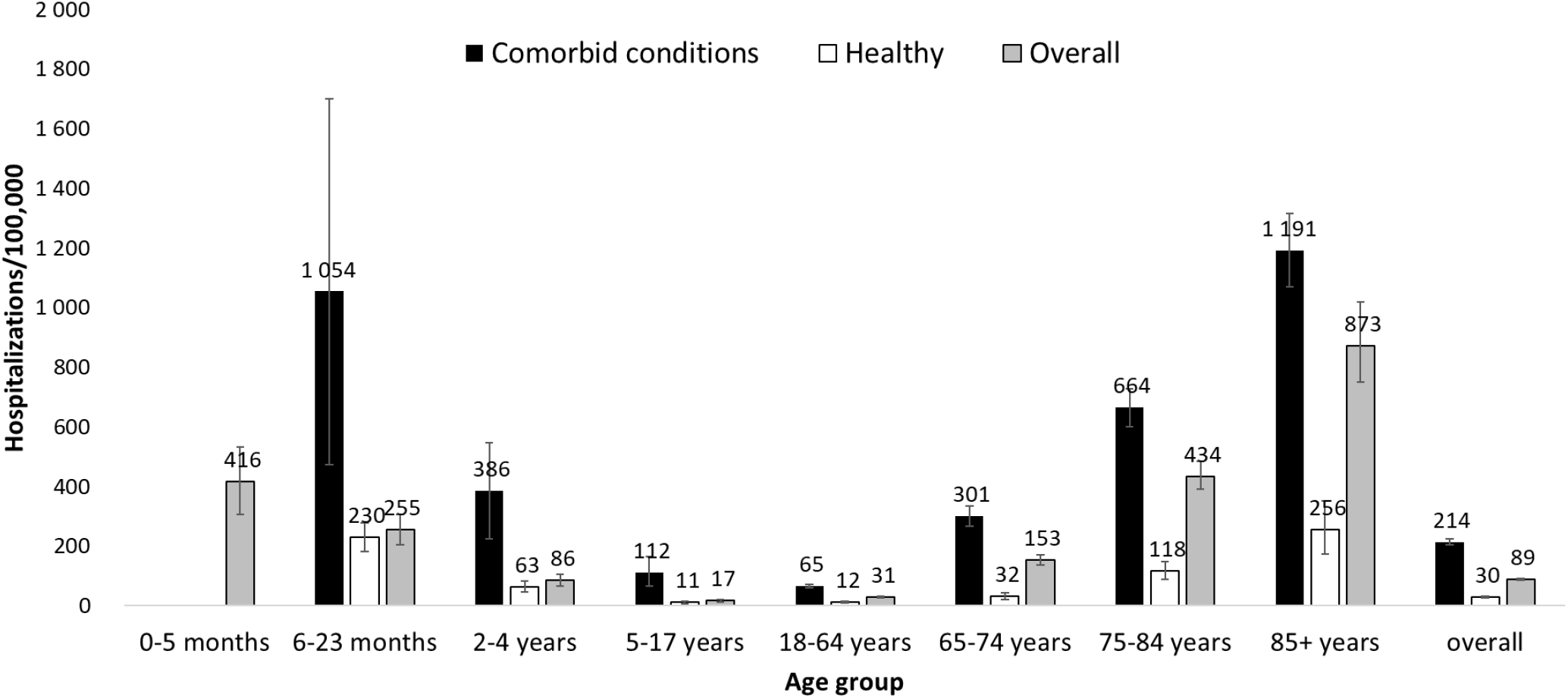
Estimated influenza-associated hospitalization rates (per 100,000) and confidence intervals, by age group and comorbidity status, Quebec hospital surveillance (2012-13 to 2018-19)

During A(H3N2) seasons, patients aged ≥75 years were the most affected, followed by infants 0-5 months, while during A(H1N1) seasons children <2 years of age had the highest hospitalization risks (**Figure 2**). For all examined seasons, hospitalization incidences were higher in patients 6-23 months and 18-64 years during A(H1N1) versus A(H3N2) seasons.

Hospitalization risks were higher for unvaccinated individuals in all adult ages both in A(H1N1) and A(H3N2) seasons and regardless of comorbidity status. The only exception was among 18-64-year-olds with comorbidity, for whom hospitalization incidences were unexpectedly higher in vaccinated individuals (**Figure 4, Supplementary Table 4**). Overall, the ratio of hospitalization risk comparing unvaccinated to vaccinated adults was higher among those without comorbidities (ranging from 2.3 for 18-64-year-olds to 6.4 for ≥85-year-olds) than among those with comorbidities (from 0.7 to 1.8 respectively) (**Supplementary Table 4**).

**Figure 4.**
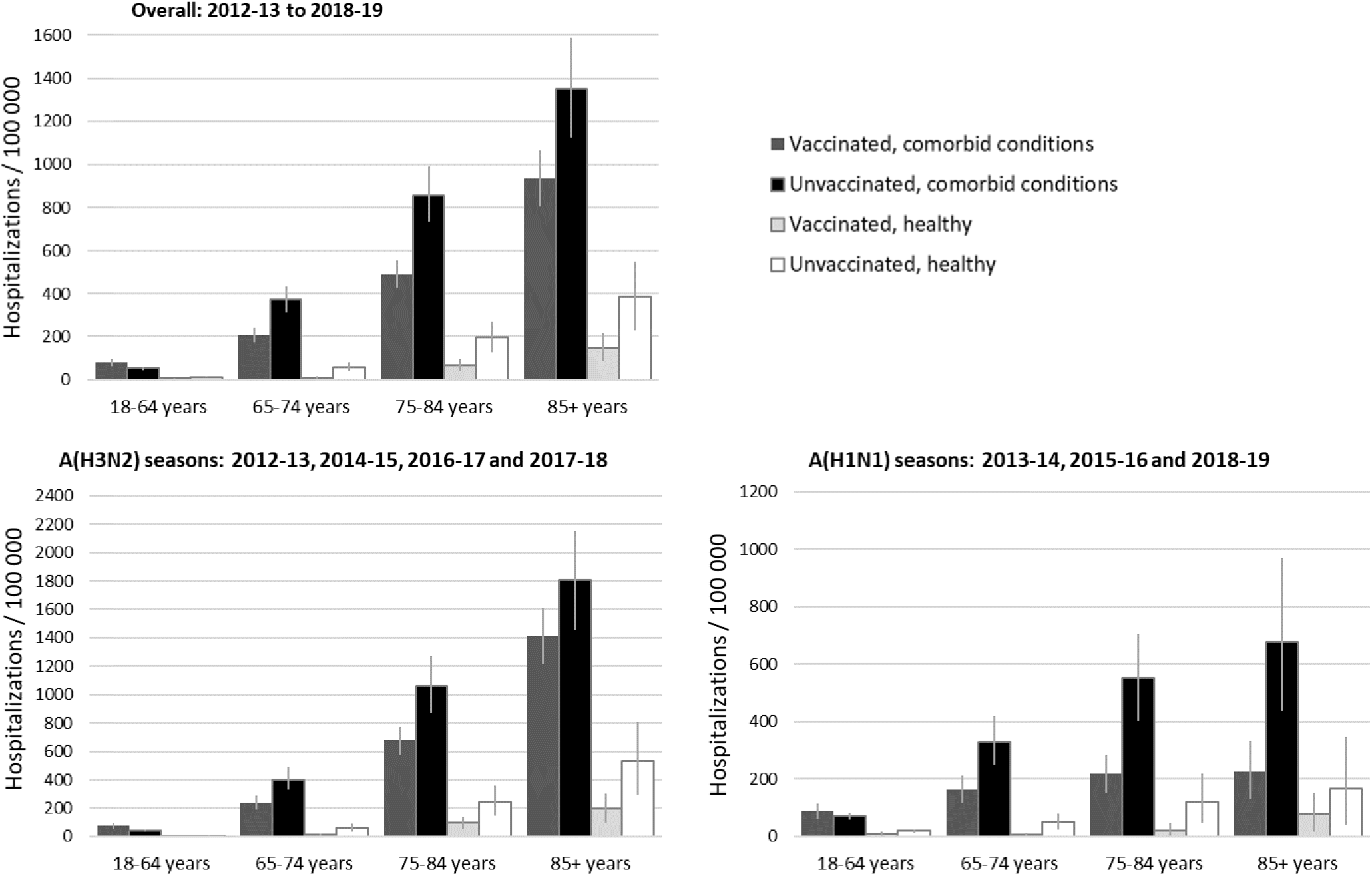
Estimated influenza-associated annual hospitalization rates (per 100,000) by vaccination status among adults with or without comorbidity, Quebec, 2012-13 to 2018-19 Note: Vertical lines in the bars represent the 95% confidence intervals

### Number needed to vaccinate

The proportion of unvaccinated cases and the NNV per hospitalization prevented varied by age and comorbidity status (**Table 3**). Individuals ≥75 years with comorbidity represented 39% of all hospitalizations; 41% were unvaccinated and the NNV (assuming VE of 50%) to prevent hospitalization was 1,995. In contrast, healthy adults 18-64 years comprised 6% of hospitalizations; 88% were unvaccinated, and the NNV to prevent hospitalization was 163,488 (82 times higher). Healthy adults 65- 74 years and those 18-64 years with comorbidity had similar NNV (respectively 35,418 vs 41,025) but the former comprised 2% of hospitalizations compared to 16% for the latter and with 80% vs 67% unvaccinated, respectively. Overall patterns were the same in sensitivity analysis ranging VE estimates from 30-60%.

**Table 3.** Number needed to vaccinate to prevent one hospitalization by age group and comorbidity status, Quebec hospital surveillance from 2012-13 to 2018-19.

| Comorbidity status and age group | % of all hospitalizations | % unvaccinated<sup>a</sup> (partly preventable) | Incidence in unvaccinated (per 100,000) | NNV to prevent one hospitalization<sup>b</sup> assuming different vaccine effectiveness |  |  |
| --- | --- | --- | --- | --- | --- | --- |
|  |  |  |  | VE=30% | VE=50% | VE=60% |
| All patients |  |  |  |  |  |  |
| 0-5 months | 2.6% | NA | NA | NA | NA | NA |
| 6-23 months | 5.0% | 79.2% | NE | NE | NE | NE |
| 2-4 years | 3.5% | 94.2% | NE | NE | NE | NE |
| 5-17 years | 2.9% | 84.9% | NE | NE | NE | NE |
| 18-64 years | 23.3% | 72.5% | 2.5 | 134,632 | 80,779 | 67,316 |
| 65-74 years | 17.5% | 56.9% | 18.6 | 17,881 | 10,729 | 8,941 |
| 75-84 years | 24.7% | 43.7% | 57.2 | 5,831 | 3,499 | 2,916 |
| ≥85 years | 20.5% | 40.2% | 97.7 | 3,411 | 2,047 | 1,706 |
| ≥65 years | 62.1% | 46.2% | 36.9 | 9,039 | 5,424 | 4,520 |
| ≥75 years | 44.8% | 42.1% | 63.4 | 5,259 | 3,155 | 2,629 |
| Patients with comorbidity |  |  |  |  |  |  |
| 0-5 months | NA | NA | NA | NA | NA | NA |
| 6-23 months | 0.7% | 58.3% | NE | NE | NE | NE |
| 2-4 years | 1.2% | 95.2% | NE | NE | NE | NE |
| 5-17 years | 1.1% | 70.0% | NE | NE | NE | NE |
| 18-64 years | 16.4% | 66.7% | 4.9 | 68,375 | 41,025 | 34,187 |
| 65-74 years | 15.1% | 53.6% | 34.5 | 9,655 | 5,793 | 4,828 |
| 75-84 years | 21.6% | 42.0% | 54.8 | 3,932 | 2,359 | 1,966 |
| ≥85 years | 17.8% | 38.7% | 134.3 | 2,482 | 1,489 | 1,241 |
| ≥65 years | 54.5% | 44.1% | 62.4 | 5,339 | 3,203 | 2,670 |
| ≥75 years | 39.4% | 40.5% | 100.2 | 3,325 | 1,995 | 1,663 |
| Patients without comorbidity |  |  |  |  |  |  |
| 0-5 months | NA | NA | NA | NA | NA | NA |
| 6-23 months | 4.7% | 82.1% | NE | NE | NE | NE |
| 2-4 years | 2.7% | 93.8% | NE | NE | NE | NE |
| 5-17 years | 1.9% | 93.9% | NE | NE | NE | NE |
| 18-64 years | 6.3% | 87.5% | 1.2 | 272,479 | 163,488 | 136,240 |
| 65-74 years | 2.2% | 79.5% | 5.6 | 59,030 | 35,418 | 29,515 |
| 75-84 years | 3.1% | 55.6% | 20.0 | 16,703 | 10,022 | 8,351 |
| ≥85 years | 2.3% | 51.2% | 37.8 | 8,828 | 5,297 | 4,414 |
| ≥65 years | 7.6% | 61.2% | 11.3 | 29,388 | 17,633 | 14,694 |
| ≥75 years | 5.4% | 53.7% | 24.8 | 13,442 | 8,065 | 6,721 |

<sup>a</sup> Including those with unknown vaccination status (4% globally)
<sup>b</sup> Number needed to vaccinate to prevent one hospitalization using the following calculation: $NNV =$
Abbreviation: NA, non-applicable; NE, non-estimable; NNV, number needed to vaccinate; VE, vaccine effectiveness

## DISCUSSION

This hospital-based surveillance network involving systematic PCR testing during seasonal peak epidemic weeks showed important differences in influenza hospitalization risk by subtype, age, comorbidity and vaccination status. Consistent with their influenza vaccination prioritization, individuals with comorbidity and those at the extremes of age were disproportionately affected. Despite decades of recommendations and promotion efforts focused toward elderly patients and those with comorbidity, however, a large percentage (nearly half) hospitalized with influenza were unvaccinated, with severe outcomes that should therefore still be considered at least partially preventable.

We estimated that people with comorbidity, both children and adults, were at about 5-10 times higher risk of influenza-associated hospitalization than otherwise healthy individuals. Greater risk among adults with comorbidity was also observed in the few studies from the US and United Kingdom similarly stratifying hospitalization by comorbidity(16–19). In children however, these studies reported comorbidity/healthy ratios of 0 to 0.6, corresponding to relatively higher hospitalization risks among healthy versus high-risk children <17 years(16–18). These studies, however, were conducted before the 2009 influenza pandemic and applied mathematical modelling to administrative datasets, with clinically-defined (vs. laboratory confirmed) influenza outcomes. Children with comorbidities may be under-detected in some studies relying only on secondary ICD-10 codes to identify comorbidities associated with a single, primarily respiratory, admission(16,17). In contrast, an Australian cohort study conducted between 2001 and 2010 similarly found 5-fold higher risk of influenza–associated hospitalization in children <10 years old with versus without pulmonary chronic diseases, ranging between 2-11-fold depending upon the underlying disease and pediatric age group(20).

Age and influenza type/subtype also seem to interact to predict influenza hospitalization burden, with higher rates among older adults during influenza A(H3N2) predominant seasons(4,16,17,21–25).

Although many influenza immunization programs have targeted adults ≥65 years, we show further age-related differences with finer examination of older adults. In fact, compared to 65-74-year-olds, hospitalization incidences were 1.4 to 3.9 times higher in 75-84-year-olds and 2.2 to 8.6 times higher in ≥85-year-olds, more pronounced during A(H3N2) seasons. Moreover, healthy 65-74-year-old adults had lower hospitalization rates than 18-64-year-old adults with comorbidity. The US Influenza Hospitalization Surveillance Network reported similar ratios (2.2 to 6.4) when comparing the same age groups, also with greater differences during A(H3N2) seasons, but did not further stratify by presence of comorbidities(22,26). Similar to our findings, other studies displaying open-ended age categories ≥75 or ≥80 years showed 2-3-fold higher hospitalization incidences among individuals with comorbidity(16,23,27).

In our study, infants 0-5 months of age had higher hospitalization risks than children aged 6-23 months (1.6-fold) or 2-4 years (4.8-fold). These ratios are lower than reported in Finnish and US studies comparing infants 0-5 months to children 6-23 months (2-fold) or 2-4 years (ranging 8-15-fold)(28–31,23). Different from older adults, hospitalization risks in our study among children <5 years were higher in A(H1N1) predominant seasons (110 to 234/100,000) than A(H3N2) predominant seasons (85 to 166/100,000). Boddington et al also reported higher hospitalization rates (rate ratio 1.2) during A(H1N1) versus A(H3N2) predominant seasons based on two A(H1N1) predominant seasons between 2010/11 through 2014/15(4). The reason for differences in susceptibility and severe outcome risk by age and subtype is not clearly understood, but birth cohort effects determined by original childhood priming history have been postulated(32).

Of note in the above comparisons, studies published elsewhere did not account for vaccination status, with coverages that may differ by region, age group, and comorbidity status also likely contributing to observed differences in estimated hospitalization risks(33). Although our study was not designed to evaluate vaccine effectiveness against hospitalization, the lower incidence of hospitalization in vaccinated compared to unvaccinated individuals likely reflects, at least in part, the protective effects (risk reductions) associated with influenza vaccination. When stratifying for age, comorbidity and influenza type/subtype season, we observed lower hospitalization incidences among vaccinated compared to unvaccinated individuals in all strata except 18-64-year-olds with comorbidity. Not all comorbidities are expected to influence VE equally and this unexpectedly higher risk among vaccinated versus unvaccinated 18-64-year-olds with comorbidity may be partially explained by a higher percentage with immunosuppressive condition among vaccinated (29%) vs. unvaccinated (13%) adults 18-64 years [not displayed].

NNV derivation enables quantification of the impact of further immunization efforts, but requires accurate estimates of absolute hospitalization risks sufficiently granular to examine potential target group comparisons. Severe outcome rates have traditionally been estimated using ecologic designs and various statistical models applied to administrative and surveillance data(34,35). Validation studies comparing ecological model-based estimates to empirical measures incorporating laboratory-confirmed outcomes show the former to be unreliable in having both over- and under-estimated hospitalization rates(36). More recently, and similar to our own study, others have also used PCR-confirmed data from influenza hospitalization surveillance networks with adjustment for under-detection, population characteristics and/or extrapolation to the general population(21,37–40). Unlike our study, however, estimates elsewhere did not take into account vaccination status(21,37–40). Additional stratification on vaccine status supports NNV estimation and points to where improvements in vaccination coverage may have the greatest and most efficient impact on residual hospitalization burden. While actual NNV estimates will vary between jurisdictions, all jurisdictions would benefit from this more granular and goal-oriented monitoring approach. Understanding the distribution of influenza hospitalizations simultaneously by age, comorbidity and vaccination status reveals the proportion of preventable and non-preventable hospitalizations by relevant target groups, and the immunization gaps to be addressed to further most efficiently reduce healthcare burden.

Our study has limitations. Sample size was limited to ≤1000 participants admitted annually across four hospitals and surveillance was conducted only during seasonal peak weeks. Consequently, we missed a large proportion of influenza B infections in the 2012-13, 2013-14, 2014-15, 2016-17 seasons.

Extrapolation of the age-distribution during peak weeks to the entire season may have biased age-specific estimates if the age distribution of influenza B cases or other influenza A sub-types differed outside these periods. Previous studies have shown higher hospitalization rates in children due to influenza B versus A(H3N2), suggesting we may have underestimated risk in younger age groups(4). However, this would not affect the specific A(H1N1) and A(H3N2) risk comparisons. There are uncertainties in the underlying assumptions used in adjusting influenza-associated hospitalization tallies and population denominators for each stratum. Although this may affect the precision of estimates, the comparison between groups (for instance, with comorbidity versus healthy) would likely remain valid. Despite these limitations, our systematic PCR testing of all patients presenting with ARI enhances current understanding of the influenza burden compared to other studies. For example, in the US Influenza Hospitalization Surveillance Network patients are tested by PCR or non-PCR tests at physician discretion requiring adjustment for the frequency of testing and diagnostic assay sensitivity and with potential bias based on physician preferences(37). Unlike previous publications based on secondary use of administrative data assembled for a different original purpose, our trained research nurses directly collected and verified information about comorbidity and vaccination status, thus minimizing misclassification bias(16–19).

In conclusion, influenza hospitalization risks vary substantially by subtype, age, comorbidity, and vaccination status. In the context of broad-based influenza immunization programs—targeted or universal—severe outcome surveillance should be stratified simultaneously by each of these factors to enable reliable and informative NNV derivation and comparison. Only then can policymakers derive a true sense of how further vaccination efforts and expenditures in their regions should be prioritized for optimal impact and efficiency.

## ACKNOWLEDGMENTS

We would like to thank Manale Ouakki (Institut national de santé publique du Québec) for her statistical support to perform the bootstrap estimations and France Bouchard (Centre de Recherche du CHU de Québec – Université Laval) for her work as coordinator of Quebec hospital–based surveillance network.**Financial support**. This work was supported by the Ministère de la santé et des services sociaux du Québec. **Author contributions.** All authors had full access to all of the data in the study and take responsibility for the integrity of the data and the accuracy of the data analysis. **Concept and design**: SC, CAG, DS, GDS, RG. **Acquisition, analysis, or interpretation of data**: All authors. **Drafting of the manuscript**: SC, CAG, RG. **Critical revision of the manuscript for important intellectual content**: All authors. **Statistical analysis**: SC, CAG. **Supervision**: GDS, RG

## Data availability

Individual data not publicly available

