## Supplemental material for "Influenza hospitalization burden by subtype, age, comorbidity and vaccination status: 2012/13 to 2018/19 seasons, Quebec, Canada"

**SUPPLEMENTARY MATERIAL**

**Supplementary Figure 1**. Proportion of each influenza type/subtype identified in participants of the Quebec hospital surveillance network during the surveillance periods and Quebec influenza type circulation according to the provincial sentinel laboratory surveillance……………………………………………………2

**Supplementary Table 1**. Detailed adjustments to estimate hospitalization incidence in the study population according to health status……………………………………………………………………………………..………..………3

**Supplementary Table 2**. Reasons for exclusion amongst eligible patients and percent participation…………4

**Supplementary Table 3**: Age-distribution and vaccination status of comorbidities (non-exclusive categories) among hospitalizations in Quebec, 2012-13 to 2018-19……………………………………………………………5

**Supplementary Table 4**: Estimated influenza-associated hospitalization rates (per 100,000) by vaccination status among adults with or without comorbidities, Quebec 2012-13 to 2018-19………………..…….6

**Supplementary Figure 1**. Proportion of each influenza type/subtype identified in participants of the Quebec hospital surveillance network during the surveillance periods and Quebec influenza type circulation according to the provincial sentinel laboratory surveillance

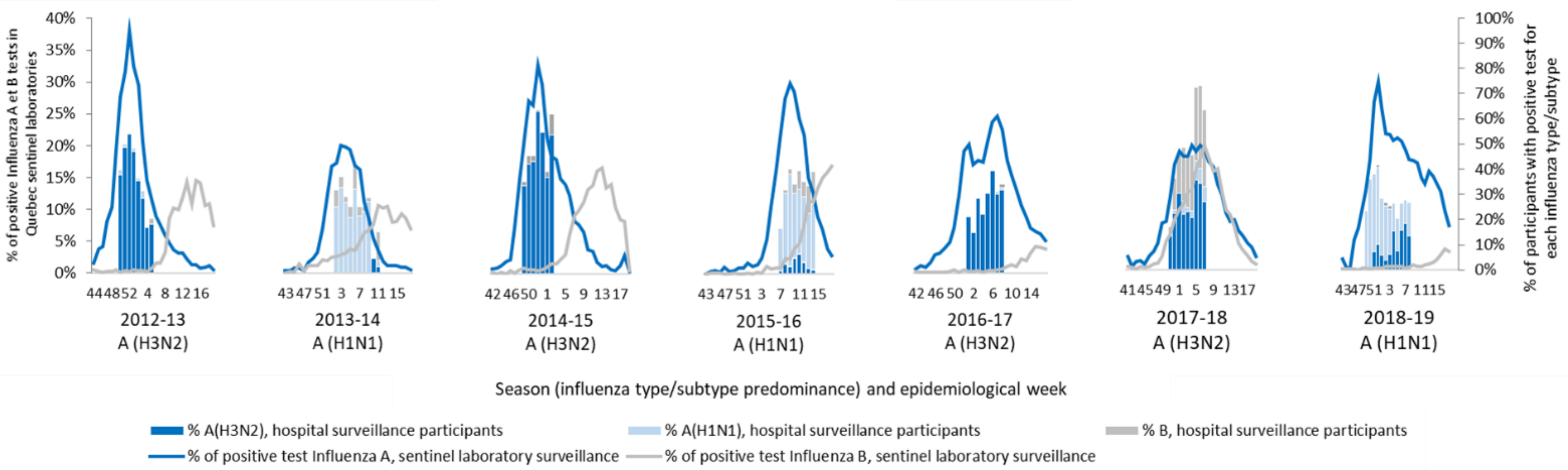


Note: Periods of the surveillance : December 9, 2012 to February 2, 2013 (epi-weeks 2012.50 to 2013.05); January 1, 2014 to March 15, 2014 (epi-weeks 2014.02 to 2014.11); November 30, 2014 to January 13, 2015 (epi-weeks 2014.49 to 2015.02); February 02, 2016 to April 04, 2016 (epi-weeks 2016.07 to 2016.14); January 1,2017 to February 25,2017 (epi-weeks 2017.01 to 2017.08); December 17, 2017 to February 17, 2018 (epi-weeks 2017.51 to 2018.07); December 09, 2018 to March 02, 2019 (epi-weeks 2018.50 to 2019.09).

**Supplementary Table 1**. Detailed adjustments to estimate hospitalization incidence in the study population according to health status

| Season | Healthy | Comorbidity |
|---|---|---|
| **1. Adjustment for under-detection[a]** | | |
| **2012-13** | x1.09 | x1.17 |
| **2013-14** | x1.10 | x1.16 |
| **2014-15** | x1.05 | x1.11 |
| **2015-16** | x1.12 | x1.15 |
| **2016-17** | x1.09 | x1.10 |
| **2017-18** | x1.03 | x1.10 |
| **2018-19** | x1.16 | x1.16 |
| **2. Adjustment for non-participation of eligible patients[b]** | | |
| **2012-13** | x1.70 | x1.70 |
| **2013-14** | x1.39 | x1.39 |
| **2014-15** | x1.40 | x1.40 |
| **2015-16** | x1.59 | x1.59 |
| **2016-17** | x1.22 | x1.22 |
| **2017-18** | x1.23 | x1.23 |
| **2018-19** | x1.15 | x1.15 |
| **3. Extrapolation to the entire season[c]** | | |
| **2012-13** | x1.37 | x1.37 |
| **2013-14** | x1.37 | x1.37 |
| **2014-15** | x1.89 | x1.89 |
| **2015-16** | x1.40 | x.140 |
| **2016-17** | x1.73 | x1.73 |
| **2017-18** | x1.62 | x1.62 |
| **2018-19** | x1.46 | x1.46 |

[a] Adjustment for under-detection due to low test sensibility among those arriving >8 days after symptoms onset. Multiplier = 1 / (proportion of detected influenza among all potential influenza cases)

[b] Adjustment for non-participation of eligible patients. Multiplier = 1 / (percentage of eligible patients who gave consent among all eligible patients)

[c] Extrapolation to the entire season according to data from the provincial sentinel laboratory surveillance. Multiplier = 1 / (proportion of positive samples during the weeks included in the surveillance among all positive samples throughout the season)

Note: Adjusted hospitalization incidences are obtained with the following equation: Adjusted incidence = measured incidence x multiplier for under-detection x multiplier for non-participation of eligible patients x multiplier for extrapolation to the entire season

**Supplementary Table 2**. Reasons for exclusion amongst eligible patients (n=6,545) and percent participation

| | Excluded | Included | Participation |
|---|---|---|---|
| | N (%) | N (%) | (%) |
| **Reasons for exclusion**: | 1,724 | 4,821 | 73.7 |
| - Refused to participate | 111 (6.4) | | |
| - Missed by study nurse | 763 (44.3) | | |
| - Unable to consent | 556 (32.3) | | |
| - Sampling/lab problem | 141 (8.2) | | |
| - Other exclusions | 153 (8.9) | | |
| **By season**: | | | |
| 2012-13 | 379 (22.0) | 542 (11.2) | 58.8 |
| 2013-14 | 215 (12.5) | 558 (11.6) | 72.2 |
| 2014-15 | 281 (16.3) | 706 (14.6) | 71.5 |
| 2015-16 | 365 (21.2) | 620 (12.9) | 62.9 |
| 2016-17 | 177 (10.3) | 791 (16.4) | 81.7 |
| 2017-18 | 177 (10.3) | 755 (15.7) | 81.0 |
| 2018-19 | 130 (7.5) | 849 (17.6) | 86.7 |

Note: Eligible patients were those hospitalized more than 24h due to a community-acquired acute respiratory infection or fever of unknown origin

**Supplementary Table 3**: Age-distribution and vaccination status of comorbidities (non-exclusive categories) among hospitalizations in Quebec, 2012-13 to 2018-19

| **Children** | **0-5 months** | | **6-23 months** | | **2-4 years** | | **5-17 years** | |
|---|---|---|---|---|---|---|---|---|
| **COMORBIDITY** | N (%) | %VACC | N (%) | %VACC | N (%) | %VACC | N (%) | %VACC |
| Heart disease | 1 (100) | 0.0 | 0 | NA | 0 | NA | 0 | NA |
| Respiratory disease | 0 | NA | 11 (91.7) | 36.4 | 19 (90.5) | 5.3 | 18 (90) | 27.8 |
| COPD | 0 | NA | 0 | NA | 0 | NA | 1 (5.0) | 100.0 |
| Asthma | 0 | NA | 11 (91.7) | 36.4 | 19 (90.5) | 5.3 | 17 (85.0) | 23.5 |
| Obesity | 0 | NA | 0 | NA | 0 | NA | 0 | NA |
| Nephropathy | 0 | NA | 1 (8.3) | 100.0 | 1 (4.8) | 0.0 | 0 | NA |
| Immunopathy | 0 | NA | 0 | NA | 0 | NA | 1 (5.0) | 100.0 |
| Neuropathy | 0 | NA | 0 | NA | 3 (14.3) | 0.0 | 1 (5.0) | 0.0 |
| Diabetes | 0 | NA | 0 | NA | 0 | NA | 0 | NA |
| Any comorbidity | 1 (100) | 0.0 | 12 (100) | 41.7 | 21 (100) | 4.8 | 20 (100) | 30.0 |
| **Adults** | **18-64 years** | | **65-74 years** | | **75-84 years** | | **85+ years** | |
| **COMORBIDITY** | N (%) | %VACC | N (%) | %VACC | N (%) | %VACC | N (%) | %VACC |
| Heart disease | 74 (24.4) | 39.2 | 145 (54.3) | 49.0 | 232 (60.6) | 60.8 | 324 (74.3) | 61.1 |
| Respiratory disease | 173 (59.5) | 34.1 | 158 (59.2) | 51.3 | 202 (52.7) | 63.9 | 147 (46.7) | 66.0 |
| COPD | 102 (35.1) | 39.2 | 130 (48.7) | 54.6 | 164 (42.8) | 67.1 | 121 (38.4) | 65.3 |
| Asthma | 111 (38.1) | 34.2 | 63 (23.6) | 46.0 | 92 (24.0) | 62.0 | 57 (18.1) | 73.7 |
| Obesity | 43 (14.8) | 27.9 | 28 (10.5) | 46.4 | 24 (6.3) | 50.0 | 6 (1.9) | 83.3 |
| Nephropathy | 31 (10.7) | 45.2 | 45 (16.9) | 44.4 | 101 (26.4) | 60.4 | 110 (34.9) | 60.0 |
| Immunopathy | 53 (18.2) | 52.8 | 51 (19.1) | 45.1 | 28 (7.3) | 53.6 | 22 (7.0) | 68.2 |
| Neuropathy | 20 (5.9) | 25.0 | 12 (4.5) | 33.3 | 24 (6.3) | 62.5 | 29 (9.2) | 58.6 |
| Diabetes | 94 (32.3) | 37.2 | 114 (42.7) | 44.7 | 151 (39.4) | 57.6 | 91 (28.9) | 55.0 |
| Any comorbidity | 291 (100) | 33.3 | 267 (100) | 46.4 | 383 (100) | 58.0 | 315 (100) | 61.3 |

Abbreviations: COPD, chronic obstructive pulmonary disease or other chronic pulmonary disease except asthma (children); NA, not applicable; VACC, vaccinated

**Supplementary Table 4**: Estimated influenza-associated hospitalization incidence (per 100,000) by vaccination status among adults with or without comorbidities, Quebec 2012-13 to 2018-19

| | | All seasons | | | A(H3N2) seasons | | | A(H1N1) seasons | | |
|---|---|---|---|---|---|---|---|---|---|---|
| **Age (years)** | **Comorbid status** | UNVACC | VACC | Ratio U/V | UNVACC | VACC | Ratio U/V | UNVACC | VACC | Ratio U/V |
| 18-64 | Comorbidity | 53.1 | 81.1 | 0.7 | 40.3 | 75.2 | 0.5 | 71.4 | 89.4 | 0.8 |
| | Healthy | 13.2 | 5.9 | 2.3 | 8.6 | 3.4 | 2.6 | 19.9 | 9.5 | 2.1 |
| 65-74 | Comorbidity | 373.0 | 206.4 | 1.8 | 403.3 | 237.9 | 1.7 | 331.7 | 163.5 | 2.0 |
| | Healthy | 58.3 | 9.2 | 6.4 | 63.5 | 11.8 | 5.4 | 51.2 | 5.6 | 9.2 |
| 75-84 | Comorbidity | 856.0 | 491.4 | 1.7 | 1064.2 | 678.6 | 1.6 | 552.5 | 218.5 | 2.5 |
| | Healthy | 196.3 | 66.2 | 3.0 | 247.7 | 96.6 | 2.6 | 121.3 | 22.0 | 5.5 |
| 85+ | Comorbidity | 1349.9 | 934.3 | 1.4 | 1804.3 | 1412.9 | 1.3 | 677.2 | 226.0 | 3.0 |
| | Healthy | 387.6 | 148.0 | 2.6 | 536.4 | 194.2 | 2.8 | 167.3 | 79.7 | 2.1 |

Abbreviations: UNVACC, unvaccinated; VACC, vaccinated; Ratio U/V, Ratio of rates among unvaccinated and rates among vaccinated
